# Early Valve Replacement vs Surveillance in Moderate Aortic Stenosis: Meta-Analysis of Randomized and Observational Evidence

**DOI:** 10.64898/2026.11.04.26367266

**Authors:** Ali Majed, Pia Kmeid, Ayah Safa, Hamza Kahwaji, Hashem Safieddine, Georges Ghanem

## Abstract

**BACKGROUND:** Moderate aortic stenosis (AS) is associated with excess mortality, and 2 randomized trials have tested early transcatheter aortic valve replacement (TAVR).

**OBJECTIVES:** The authors sought to estimate the effects of early TAVR vs clinical surveillance and to compare randomized with observational estimates.

**METHODS:** Six databases and registries were searched through November 3, 2026. Adjusted observational studies were synthesized separately from trials. Certainty was rated with GRADE.

**RESULTS:** Two trials (922 patients; TAVR UNLOAD, n = 178, HF with reduced ejection fraction; PROGRESS, n = 744, 96.4% symptomatic) and 7 cohorts were included. Early TAVR did not detectably change death over approximately 2 years (HR: 1.05; 95% CI: 0.74-1.49; low certainty), with one-fifth of the deaths needed to detect a 20% reduction. Heart failure (HF) hospitalization was lower (RR: 0.53; 95% CI: 0.36-0.77; low certainty), or 70 fewer patients per 1,000 at 2 years. New pacemakers were more frequent (PROGRESS: 17.3% vs 9.3%; moderate certainty). About half of surveillance patients underwent valve replacement within 2 years. Observational cohorts estimated a 48% mortality reduction (HR: 0.52; 95% CI: 0.43-0.64; very low certainty), which differed from the randomized estimate (ratio of HRs: 2.01; 95% CI: 1.37-2.95).

**CONCLUSIONS:** In older, predominantly symptomatic patients with moderate AS, early TAVR may make little or no difference to 2-year mortality and may reduce HF hospitalization, at the cost of more pacemakers. Because about half of patients under surveillance receive a valve within 2 years, the decision largely concerns timing. Observational survival estimates should not guide practice.

## Introduction

Guidelines reserve aortic valve replacement (AVR) for severe aortic stenosis (AS) and recommend surveillance every 1 to 2 years for moderate disease.^1,2^ Yet mortality rises well below severe thresholds,^3^ reaching about 9 deaths per 100 person-years in moderate AS,^4^ and is high when moderate AS accompanies left ventricular dysfunction.^5^ Early AVR reduced hospitalization in asymptomatic severe AS,^6,7^ making moderate AS with symptoms or cardiac damage the next population to test.

Two randomized trials have now reported. TAVR UNLOAD randomized 178 patients with heart failure with reduced ejection fraction (HFrEF) and did not show superiority on a hierarchical endpoint, although health status improved.^8^ PROGRESS randomized 744 patients with at-risk features and reported fewer deaths or heart failure (HF) events with TAVR.^9,10^ The trials differ in phenotype, in how HF events were counted, and in the extent of crossover to AVR. The observational studies that motivated them reported survival benefits of about 50%,^11,12^ and randomized evidence can now test those estimates.

We therefore pooled only outcomes that both trials defined in common, set the benefit against procedural harm in absolute terms, examined whether the effect depends on phenotype or cardiac damage, and formally compared the randomized estimate with that of adjusted observational studies.

## Methods

### Protocol and registration

The review followed PRISMA 2020^13^ (Supplemental Table 1). The protocol was drafted before PROGRESS was presented and registered with PROSPERO (CRD420261559767) the next day, November 3, 2026, before screening began. Ethical approval was not required because only published data were analyzed. Deviations are listed in Supplemental Table 2; analyses they introduced, including the harmonization of HF hospitalization and numerical Bayesian priors, are post hoc.

### Search and eligibility

MEDLINE, Embase, and ClinicalTrials.gov were searched from inception to November 3, 2026, without language restriction, supplemented by Web of Science, CENTRAL, the WHO ICTRP portal, and backward and forward citation searching (Supplemental Methods, Supplemental Table 3). Eligible studies enrolled adults with moderate AS as defined by each study and compared early AVR, transcatheter or surgical, with clinical surveillance in which AVR was permitted on progression. Randomized trials formed the comparative tier. Observational studies with adjusted estimates formed a separate tier, never pooled with trials; when several estimates were reported, we used the one best matching early AVR. Unadjusted comparisons and cohorts in which AVR followed progression to severe AS were excluded.

### Selection and data extraction

Two reviewers (A.M. and P.K.) independently screened records, assessed full texts, extracted data, and judged risk of bias; a third (G.G.) resolved disagreements. Agreement on screening was substantial (κ = 0.80; 95% CI: 0.64-0.96), and extracted values agreed for every study. For TAVR UNLOAD, the cause and day of each death came from the per-patient listing, allowing a common 2-year horizon; PROGRESS safety outcomes and subgroup estimates came from its supplement and forest plot.

### Outcomes and harmonization

The primary outcomes were all-cause death and HF hospitalization, pooled only as hospitalization: the first disease-related event at 1 year in TAVR UNLOAD and the first HF hospitalization at 2 years in PROGRESS. Each trial’s broader HF event outcome was pooled as a secondary outcome, and the TAVR UNLOAD hierarchical endpoint was reported as published.^14^ Other secondary outcomes were cardiovascular and non-cardiovascular death, stroke, health status, PROGRESS safety outcomes by assigned arm,^15,16^ and periprocedural complications of every AVR in either arm (Supplemental Table 4).

### Risk of bias and certainty

Trials were assessed with RoB 2 separately for death and for HF outcomes,^17^ and observational studies with ROBINS-I, with attention to confounding by indication and immortal time.^18,19^ Certainty was rated with GRADE for each tier separately.^20^ Imprecision was judged against thresholds of appreciable benefit and harm and against the optimal information size.^21^

### Statistical analysis

With 2 trials, between-trial variance cannot be estimated reliably, so the primary model was common-effect (inverse variance for log HRs; Mantel-Haenszel for RRs). Random-effects models (restricted maximum likelihood with the modified Hartung-Knapp-Sidik-Jonkman adjustment) were sensitivity analyses;^22^ IZ was reported but not used to decide whether to pool.^23^ Absolute effects at 2 years applied the pooled relative effect to the PROGRESS surveillance-arm risk and, for death, also to the TAVR UNLOAD risk; numbers needed to treat or harm were rounded up. The optimal information size was the number of events needed to detect a 20% to 25% relative reduction with 80% power and a 2-sided α of 0.05, and trial sequential analysis with O’Brien-Fleming monitoring boundaries tested whether cumulative evidence was conclusive.^24^ A Bayesian random-effects model (normal prior on the log HR with mean 0 and SD 1; half-normal prior on τ with scale 0.5, and 0.25 and 1.0 in sensitivity analyses) estimated the probability of benefit.^25^ The intention-to-treat estimate was primary; crossover was tabulated, and crossover-censored estimates were reported as published.^26^

Differences between trials, between subgroups within PROGRESS, and between the randomized and observational tiers were tested as ratios of relative effects,^27^ and the credibility of effect modification was judged with ICEMAN.^28^ TAVR UNLOAD did not report outcomes by sex.

Sensitivity analyses are listed in Supplemental Table 8. Observational HRs were pooled by random effects, recognizing that pooling does not remove shared confounding,^29^ and the confounding needed to explain them was quantified with the E-value.^30^ Risk of bias due to missing evidence was assessed with ROB-ME.^31^ Analyses used Python 3 (NumPy, SciPy), with 2-sided P values.

### Data sharing statement

All data were extracted from published sources and are presented in the article and Supplemental Appendix. The extraction dataset and analysis code are available at https://doi.org/10.5281/zenodo.23950111.

## Results

### Study selection and trial characteristics

The searches identified 9,383 records. After removal of 1,349 duplicates and 3,745 records excluded by keyword prefilters, 4,289 records were screened and 18 reports assessed in full, with 5 more from citation searching (Supplemental Figure 1, Supplemental Table 5). Two trials (462 early TAVR, 460 surveillance)^8,10^ and 7 adjusted observational studies^32–38^ were included. A cohort in which AVR followed progression to severe AS^39^ and 4 prognostic cohorts from an earlier meta-analysis^11^ were excluded. Three TAVR UNLOAD sub-analyses and an observational abstract were summarized but not pooled.^40–43^

The trials covered 2 phenotypes (Table 1, Supplemental Table 6). TAVR UNLOAD enrolled patients with HFrEF (mean LVEF 39%; 55.6% in NYHA functional class III or IV).

**TABLE 1.** Design, Population, and Main Outcomes of the Randomized Trials.

|  | **TAVR UNLOAD** | **PROGRESS** |
|---|---|---|
| Publication; sponsor | J Am Coll Cardiol 2025; investigator-initiated | N Engl J Med 2026; valve manufacturer |
| Phenotype | HFrEF on guideline-directed therapy, LVEF 20%-50%, NYHA II-IV | Age ≥65 y with ≥1 at-risk feature (symptoms, LVEF <60%, diastolic dysfunction, low flow, AF, NT-proBNP >3× ULN, high calcium score) |
| Definition of moderate AS | AVA >1.0 and ≤1.5 cm²; DSE in 45%; core laboratory | Core laboratory; 91.6% concordant on area and velocity or gradient |
| n (TAVR/surveillance) | 89/89 | 373/371 |
| Age, y; women | 77.4; 20.8% | 78.4; 39.1% |
| LVEF | Mean 39.0% | Mean 63.4% (<50% in 8.5%) |
| NYHA II-IV; III-IV | 98.9%; 55.6% | 96.4%; 35.5% |
| STS-PROM | 4.4% | 3.0% |
| Follow-up | Median 23 mo | 2 y |
| All-cause death | 35 vs 35; HR: 0.98 (0.61-1.56) | 30 vs 26; HR: 1.14 (0.67-1.93) |
| HF hospitalization | 10 vs 16 (first event, 1 y) | 26 vs 52 (2 y) |
| Trial-defined HF event | 34 vs 40; HR: 0.80 (0.51-1.27) | 38 vs 66; HR: 0.55 (0.37-0.81) |
| Primary endpoint | Win ratio: 1.31 (0.91-1.88) | Death or HF event: 16.7% vs 23.9%; HR: 0.67 (0.48-0.93) |
| AVR in surveillance arm | 38 (42.7%), median 12 mo | 204 (55.0%); Kaplan-Meier 57.4% at 2 y |
| Events committee | Blinding not stated | Aware of assignment |

Values are for both arms combined unless shown as TAVR vs surveillance; ratios are with 95% CIs; percentages for the PROGRESS primary endpoint are Kaplan-Meier estimates. Full baseline characteristics are in Supplemental Table 6. AF = atrial fibrillation; AS = aortic stenosis; AVA = aortic valve area; DSE = dobutamine stress echocardiography; HF = heart failure; HFrEF = heart failure with reduced ejection fraction; HR = hazard ratio; LVEF = left ventricular ejection fraction; NT-proBNP = N-terminal pro-B-type natriuretic peptide; NYHA = New York Heart Association; STS-PROM = Society of Thoracic Surgeons Predicted Risk of Mortality; TAVR = transcatheter aortic valve replacement; ULN = upper limit of normal.

PROGRESS enrolled patients aged 65 years or older with at least 1 at-risk feature; 96.4% had symptoms and 8.5% had an LVEF below 50%. Both used transfemoral balloon-expandable valves.

Both trials were at low risk of bias for death. For HF outcomes, both were at high risk in outcome measurement: PROGRESS’s events committee was aware of assignment, and TAVR UNLOAD did not state that adjudication was blinded (Supplemental Figure 2, Supplemental Table 7). All observational studies were at serious risk of bias, from confounding by indication in all and time-related bias (immortal time or misaligned time zero) in 4. Both trials reported all registered outcomes, and no completed but unpublished trial was identified.

### Primary outcomes

Early TAVR did not detectably change all-cause death (2 trials, 126 deaths; HR: 1.05; 95% CI: 0.74-1.49; P = 0.80; IZ = 0%) (Figure 1A). Truncated at a common 2-year horizon, the RR was 1.19 (95% CI: 0.83-1.71). At PROGRESS-like risk, this corresponds to 3 more deaths per 1,000 at 2 years (19 fewer to 34 more), and at TAVR UNLOAD-like risk to 9 more (54 fewer to 91 more). Detecting a 20% relative reduction with 80% power requires about 630 deaths; the trials provide one-fifth of that number, and the cumulative Z-curve (Z = 0.26) was far from any monitoring boundary.

**FIGURE 1.**
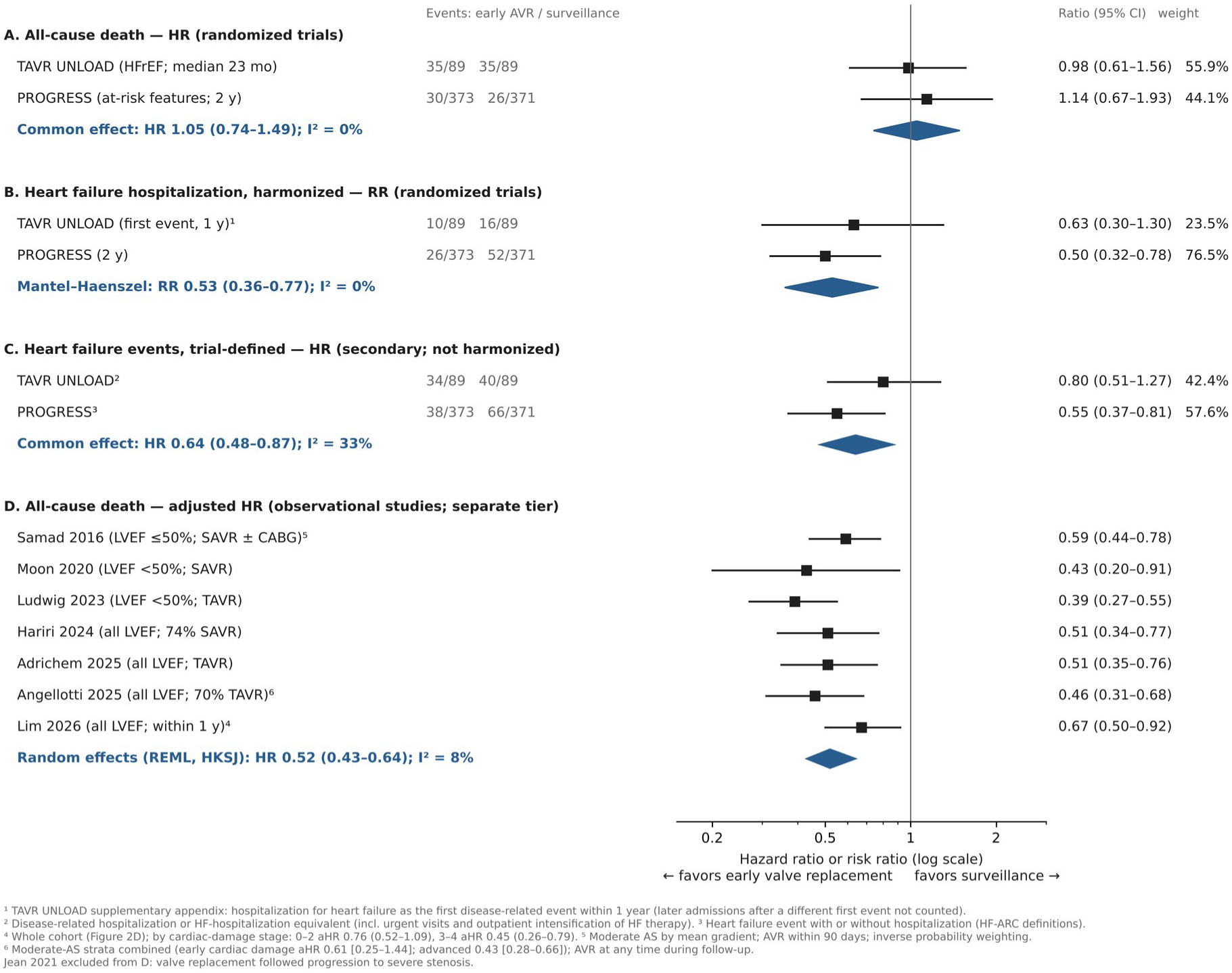
Primary Outcomes of Early TAVR vs Surveillance. Randomized trials: (A) all-cause death, (B) HF hospitalization, (C) trial-defined HF events. (D) Adjusted HRs for death from 7 observational studies, never pooled with the trials. Squares show study estimates (95% CI) sized by weight; diamonds show pooled estimates. Early TAVR reduced HF hospitalization but not death; observational studies suggested a 48% mortality reduction. AVR = aortic valve replacement; CABG = coronary artery bypass grafting; HF = heart failure; HFrEF = heart failure with reduced ejection fraction; HKSJ = Hartung-Knapp-Sidik-Jonkman; HR = hazard ratio; LVEF = left ventricular ejection fraction; REML = restricted maximum likelihood; RR = risk ratio; SAVR = surgical aortic valve replacement; TAVR = transcatheter aortic valve replacement.

HF hospitalization was lower with early TAVR (RR: 0.53; 95% CI: 0.36-0.77; P = 0.001; IZ = 0%) (Figure 1B): 26 of 373 vs 52 of 371 patients in PROGRESS at 2 years and 10 of 89 vs 16 of 89 in TAVR UNLOAD at 1 year. At PROGRESS-like risk, this is 70 fewer patients hospitalized per 1,000 at 2 years (95% CI: 95-34 fewer), a number needed to treat of 15 (95% CI: 11-30). HF hospitalization in PROGRESS lay beyond the point at which its prespecified hierarchical testing stopped, because the preceding secondary endpoint (death, stroke, or HF hospitalization: 17.7% vs 23.3%; P = 0.07) was not significant. Random-effects models widened the interval to include no effect (Bayesian RR: 0.56; 95% credible interval: 0.32-1.15; probability of a reduction 95%) (Supplemental Table 8). In trial sequential analysis, the cumulative Z for HF hospitalization (3.28) only just crossed the O’Brien-Fleming boundary (3.26) at 36% of the required information size of about 2,560 patients.

### Secondary outcomes and harms

Trial-defined HF events were reduced (HR: 0.64; 95% CI: 0.48-0.87; IZ = 33%) (Figure 1C), more clearly in PROGRESS than in TAVR UNLOAD. Cardiovascular death (RR: 0.91; 95% CI: 0.62-1.32), non-cardiovascular death at 2 years (RR: 1.59; 95% CI: 0.85-3.00), and stroke (RR: 1.00; 95% CI: 0.56-1.77) did not differ (Figure 2). In PROGRESS, new permanent pacemakers were implanted in 17.3% vs 9.3% of patients without a pacemaker at baseline (80 more per 1,000; 95% CI: 28-132 more; number needed to harm 13; 95% CI: 8-36), whereas disabling stroke, myocardial infarction, life-threatening bleeding, and new atrial fibrillation did not differ significantly. Health status improved more with early TAVR: the KCCQ score rose by 12.8 vs 3.2 points at 1 year in TAVR UNLOAD and by 14.3 vs 7.7 points at 2 years in PROGRESS (Supplemental Table 9).

**FIGURE 2.**
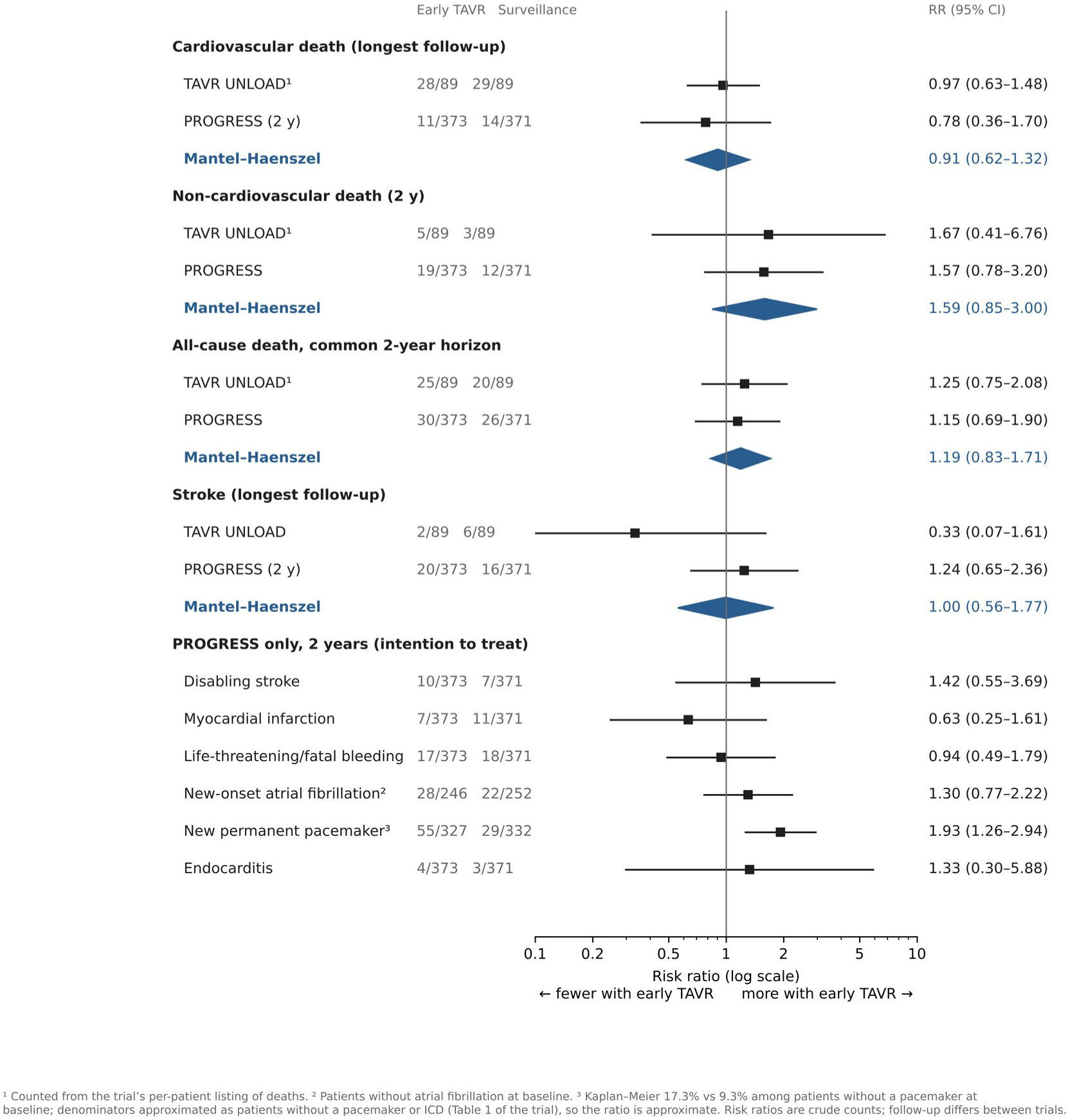
Secondary Outcomes and Harms. Risk ratios for cardiovascular death at longest follow-up, non-cardiovascular death and all-cause death at a common 2-year horizon, and stroke from both trials, and 2-year safety outcomes by assigned arm from PROGRESS. TAVR UNLOAD deaths were counted from the trial’s per-patient listing. Pacemaker estimates are among patients without a pacemaker at baseline, with approximate denominators. Squares show trial estimates and diamonds pooled Mantel-Haenszel estimates. Early TAVR increased pacemaker implantation without a detectable effect on death or stroke. ICD = implantable cardioverter-defibrillator; TAVR = transcatheter aortic valve replacement.

### Crossover and deferred replacement

During follow-up, 42.7% of surveillance patients in TAVR UNLOAD (median 12 months) and 55.0% in PROGRESS (Kaplan-Meier estimate 57.4% at 2 years) underwent AVR, 52.6% overall, mostly for progression to severe AS. Deferred replacement was often preceded by decompensation: in PROGRESS, 28 of 66 surveillance-arm HF events (42.4%) were admissions during which deferred AVR was performed, and in TAVR UNLOAD, 17 of the 35 patients who crossed over for progression had a disease-related hospitalization or equivalent first.

Descriptively, 30-day complications after deferred AVR in PROGRESS were not lower than after early TAVR (stroke 2.5% vs 0.8%; vascular complications 6.9% vs 2.0%) (Supplemental Table 10), and the KCCQ score fell by a mean of 9.3 points before deferred AVR.

### Effect modification

No difference between trials was detectable for death (ratio of HRs: 0.86; P = 0.68), HF hospitalization (ratio of RRs: 1.26; P = 0.60), or HF events (ratio of HRs: 1.45; P = 0.22). Within PROGRESS, the effect on the primary endpoint was similar with LVEF below 50% (HR: 0.71; 95% CI: 0.31-1.63; n = 63) and at least 50% (HR: 0.65; 95% CI: 0.46-0.93; interaction P = 0.85), in women (HR: 0.75; 95% CI: 0.45-1.26) and men (HR: 0.62; 95% CI: 0.40-0.95; interaction P = 0.58), and across age, symptom class, NT-proBNP, and atrial fibrillation (Figure 3, Supplemental Table 11). For cardiac damage, the HR was 0.22 (95% CI: 0.06-0.79) in stage 0-1 (16 events) and 0.75 (95% CI: 0.52-1.08) in stage 2-4 (interaction P = 0.07), 1 of 11 subgroup variables examined; credibility by ICEMAN was low. Observational cohorts reported larger effects with advanced damage, without significant interaction (Lim: P = 0.12; Angellotti: P = 0.48).^37,38^ In TAVR UNLOAD, damage stage did not modify the effect on major adverse events, but health status improved more with right-heart damage (interaction P = 0.01).^41^

**FIGURE 3.**
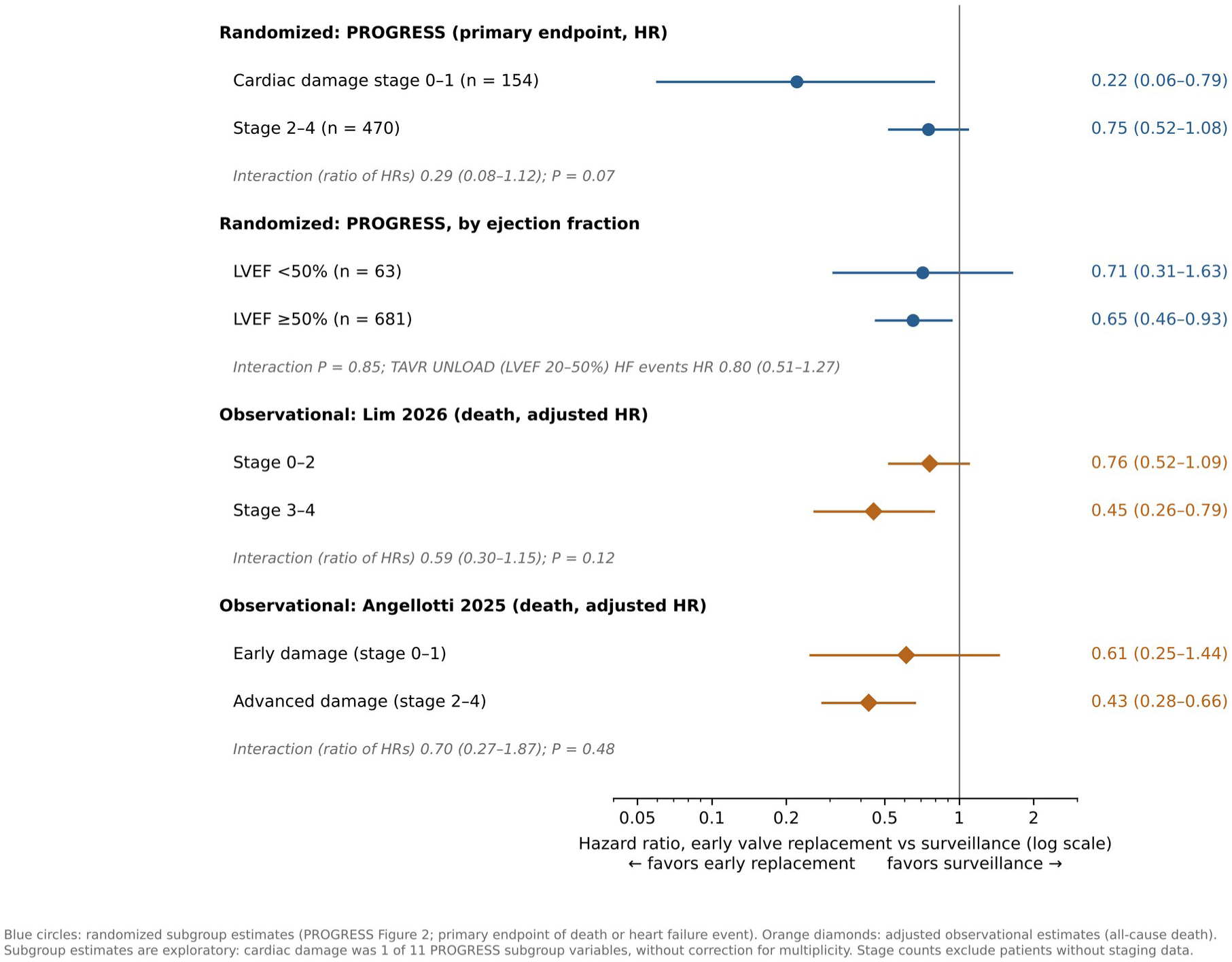
Effect Modification by Cardiac Damage and Ejection Fraction. Exploratory subgroup estimates from PROGRESS for its primary endpoint of death or HF event (circles) compared with adjusted observational estimates for all-cause death by cardiac damage stage (diamonds). In PROGRESS, point estimates favored early TAVR at least as much with stage 0-1 damage and were similar across ejection fraction; no interaction, randomized or observational, was statistically significant. HF = heart failure; HR = hazard ratio; LVEF = left ventricular ejection fraction; TAVR = transcatheter aortic valve replacement. **CENTRAL ILLUSTRATION Early Valve Replacement vs Surveillance in Moderate Aortic Stenosis** Per 1,000 patients treated early, about 70 fewer were hospitalized for HF and about 80 more received a pacemaker over 2 years, without a detectable effect on mortality. About half of surveillance patients received a valve within 2 years. The 48% mortality reduction estimated by observational studies was not reproduced by randomization. AVR = aortic valve replacement; HF = heart failure; HR = hazard ratio; KCCQ = Kansas City Cardiomyopathy Questionnaire; NNH = number needed to harm; NNT = number needed to treat; RR = risk ratio; TAVR = transcatheter aortic valve replacement.

### Observational tier

Seven adjusted observational studies estimated an HR for death of 0.52 (95% CI: 0.43-0.64; IZ = 8%) (Figure 1D, Supplemental Table 12). Estimates were stable in leave-one-out analysis (0.49-0.55), after exclusion of studies with time-related bias (0.50; 95% CI: 0.29-0.86), with concomitant surgery or pseudo-severe AS, with alternative adjusted estimates, and after inclusion of the excluded cohort or an observational abstract (0.51-0.56) (Supplemental Table 8). The observational estimate differed from the randomized one (ratio of HRs: 2.01; 95% CI: 1.37-2.95; P < 0.001). An unmeasured confounder associated with both AVR and death by an RR of 2.5 (2.1 for the CI bound) would suffice to explain it.

### Certainty of evidence

Certainty was low for mortality and HF hospitalization, moderate for pacemaker implantation, and very low for the observational mortality estimate (Table 2, Supplemental Table 13).

**TABLE 2.** GRADE Summary of Findings: Early TAVR vs Clinical Surveillance.

| **Outcome (follow-up)** | **Studies; patients** | **Relative effect (95% CI)** | **Absolute effect per 1,000 at 2 y (95% CI)** | **Certainty** | **Reasons for rating down** |
|---|---|---|---|---|---|
| All-cause death (median 23 mo to 2 y) | 2 RCTs; 922 | HR: 1.05 (0.74-1.49) | 3 more (19 fewer to 34 more) | Low | Imprecision (−2): CI includes appreciable benefit and harm; 20% of optimal information size |
| HF hospitalization (1-2 y) | 2 RCTs; 922 | RR: 0.53 (0.36-0.77) | 70 fewer (95 to 34 fewer); NNT 15 | Low | Risk of bias (−1): open label, unblinded adjudication; imprecision (−1): optimal information size not met |
| HF event, trial-defined (median 23 mo to 2 y) | 2 RCTs; 922 | HR: 0.64 (0.48-0.87) | 63 fewer (94 to 23 fewer) | Very low | Risk of bias (−1); indirectness (−1): definitions differ; imprecision (−1): 178 of 380 events needed |
| Cardiovascular death (median 23 mo to 2 y) | 2 RCTs; 922 | RR: 0.91 (0.62-1.32) | 4 fewer (14 fewer to 12 more) | Low | Imprecision (−2) |
| Stroke (median 23 mo to 2 y) | 2 RCTs; 922 | RR: 1.00 (0.56-1.77) | 0 fewer (19 fewer to 33 more) | Low | Imprecision (−2) |
| New permanent pacemaker (2 y) | 1 RCT; 744 | RR: ~1.9 (1.3-2.9) | 80 more (28 to 132 more); NNH 13 | Moderate | Indirectness (−1): 1 trial of 1 valve type |
| Health status, KCCQ (1-2 y) | 2 RCTs; 922 | Not pooled (different time points) | +9.6 points at 1 y; +6.6 at 2 y | Low | Risk of bias (−2): open label, patient-reported |
| Death, observational tier | 7 cohorts | HR: 0.52 (0.43-0.64) | — | Very low | Confounding by indication; time-related bias; inconsistent with randomized evidence |

Absolute effects apply pooled relative effects to PROGRESS surveillance-arm 2-year risks per 1,000 (death 73, HF hospitalization 149, HF event 190 [Kaplan-Meier]; cardiovascular death 38, stroke 43 [crude]); at TAVR UNLOAD-like risk (death 225), mortality is 9 more (54 fewer to 91 more). Pacemaker: PROGRESS Kaplan-Meier difference among patients without a baseline pacemaker; CI from approximate denominators. NNT and NNH are rounded up. GRADE = Grading of Recommendations Assessment, Development and Evaluation; HF = heart failure; HR = hazard ratio; KCCQ = Kansas City Cardiomyopathy Questionnaire; NNH = number needed to harm; NNT = number needed to treat; RCT = randomized controlled trial; RR = risk ratio; TAVR = transcatheter aortic valve replacement.

## Discussion

In 922 randomized patients with moderate AS, early TAVR may make little or no difference to 2-year mortality and may reduce HF hospitalization, at the cost of more permanent pacemakers (Central Illustration). Treating 1,000 PROGRESS-like patients early would prevent about 70 HF hospitalizations and add about 80 pacemakers over 2 years; the harm is estimated with more certainty than the benefit. The observational estimate that AVR halves mortality differs from the randomized one by more than chance would explain.

### Comparison with previous evidence

Previous meta-analyses of moderate AS pooled prognosis^4,44^ or observational treatment effects.^11,12^ A reconstructed-data meta-analysis restricted to LVEF ≤50% reported that AVR halved mortality (HR: 0.52; 95% CI: 0.44-0.60),^12^ yet in the same phenotype TAVR UNLOAD found no difference (HR: 0.98). The randomized pattern in moderate AS mirrors that in asymptomatic severe AS, in which a meta-analysis of 4 trials found fewer hospitalizations with early AVR but no significant mortality difference.^7^ To our knowledge, this is the first synthesis of the randomized trials in moderate AS and the first formal test of whether randomized and observational estimates agree.

### Phenotype and cardiac damage

The trials tested different hypotheses: whether relieving afterload helps a ventricle failing from another cause (TAVR UNLOAD), and whether replacing a moderately stenotic valve prevents decompensation in older, mostly symptomatic patients (PROGRESS). The HF benefit is carried by PROGRESS, in which 91.5% of patients had an LVEF of at least 50%. Pooling them estimates the effect of a strategy of early TAVR across moderate AS, the question guidelines must now answer, and tests formally showed no difference between the trials; trial-level estimates are shown throughout. TAVR UNLOAD pointed the same way but was underpowered: it was planned for 600 patients and stopped at 178. The PROGRESS subgroup with LVEF below 50% (n = 63) is too small to inform HFrEF, so evidence in that phenotype rests on TAVR UNLOAD and is inconclusive.

The observational literature suggested that early AVR helps most when extravalvular damage is advanced.^37,38^ Neither trial supported this for clinical events: in PROGRESS, point estimates favored early TAVR at least as much in patients with stage 0-1 damage. With 16 events, a nonsignificant interaction, and 11 variables examined, this may reflect chance, but it argues against restricting early AVR to advanced damage.

### Why observational and randomized estimates disagree

Three mechanisms are visible in the cohorts themselves. First, confounding by indication: patients offered AVR are those a heart team judges fit to benefit; in 1 cohort the AVR group was 8 years younger,^33^ and in another adjustment alone moved the HR from 0.92 to 0.51.^36^ Second, time-related bias: in 4 of 7 cohorts, treated patients had to survive to receive AVR, or their follow-up started later.^19^ Third, a different question: the cohorts compared AVR with no AVR, whereas the trials compared early with deferred AVR. Their agreement (IZ = 8%) reflects shared bias, not corroboration.^29^

### A trial of timing

Because about half of surveillance patients received a valve, the trials compare replacing now with replacing on progression, and the measured benefit is the cost of waiting. Deferral was often preceded by decompensation, so part of the benefit of early TAVR is avoidance of the admission that would have prompted replacement anyway. Because deferred procedures were triggered by progression, this cannot show that deferral causes harm. The other half of surveillance patients did not need a valve within 2 years, and for them early TAVR brings forward procedural risk, including pacemaker implantation, without a benefit that the trials could measure. A crossover-censored TAVR UNLOAD analysis favored TAVR,^40^ but such censoring is informative.

### Clinical implications

For patients resembling those in PROGRESS (aged 65 years or older, predominantly symptomatic, with preserved ejection fraction, and suitable for transfemoral balloon-expandable TAVR), early TAVR may be considered when symptoms persist despite treatment of coexisting conditions and avoiding HF admission is a priority, once severe AS has been excluded, provided patients understand that it is unlikely to prolong life over 2 years, that it roughly doubles the chance of a pacemaker, and that about half of patients at this stage need a valve within 2 years anyway.^45^ Evidence in HFrEF is inconclusive; there is almost none for asymptomatic patients or patients younger than 65 years, and none for surgical AVR or self-expanding valves (Supplemental Table 14).

### Future research

Pooled individual-participant data from both trials would allow time-to-event analysis at a common horizon and a direct test of effect modification by damage stage.

### Study limitations

The randomized evidence comprises 2 open-label trials, and PROGRESS contributes 81% of patients. HF outcomes are susceptible to unblinded adjudication, and PROGRESS was sponsored by the valve manufacturer. The harmonization of HF hospitalization is approximate: TAVR UNLOAD reported only first events at 1 year, and crude proportions ignore censoring and competing death; PROGRESS alone, at 2 years, gave a similar estimate (RR: 0.50; 95% CI: 0.32-0.78). Safety outcomes by assigned arm were available only from PROGRESS, and the pacemaker estimate uses approximate denominators. Follow-up is short relative to the lifetime of a bioprosthesis, so durability, leaflet thrombosis, and reintervention cannot be compared. The protocol was registered the day after PROGRESS reported, and several analyses were specified afterward. Embase-only and Web of Science records were prefiltered by keyword before screening, which could miss studies that do not use the term moderate.

## Conclusions

In older, predominantly symptomatic patients with moderate AS, early TAVR may make little or no difference to mortality over 2 years. It may reduce HF hospitalization, chiefly on the evidence of PROGRESS, at the cost of about twice as many new pacemakers. Because about half of patients under surveillance receive a valve within 2 years, the choice largely concerns timing.

The evidence does not extend to asymptomatic patients, to surgical AVR, or to self-expanding valves. Observational estimates of a survival benefit should not guide practice.

## Data Availability

All data analyzed were extracted from published sources and are presented in the manuscript and Supplemental Appendix. The extraction dataset and analysis code are available at https://doi.org/10.5281/zenodo.23950111.

## Clinical Perspectives

**WHAT IS KNOWN?** Observational studies suggested that valve replacement halves mortality in moderate AS, and 2 randomized trials of early TAVR have now reported.

**WHAT IS NEW?** In 922 randomized patients, early TAVR reduced HF hospitalization (number needed to treat 15 over 2 years) but not 2-year mortality, and increased pacemaker implantation (number needed to harm 13); about half of patients under surveillance received a valve within 2 years. The observational survival benefit differed from the randomized estimate and should not guide the timing of valve replacement.

**WHAT IS NEXT?** Future trials should adjudicate HF events blind to assignment, ascertain late valve outcomes in both strategies, and test with individual-participant data whether early AVR benefits patients before cardiac damage is established.

## ABBREVIATIONS AND ACRONYMS

AS: aortic stenosis
AVR: aortic valve replacement
GRADE: Grading of Recommendations Assessment, Development and Evaluation
HF: heart failure
HFrEF: heart failure with reduced ejection fraction
HR: hazard ratio
KCCQ: Kansas City Cardiomyopathy Questionnaire
LVEF: left ventricular ejection fraction
RR: risk ratio
TAVR: transcatheter aortic valve replacement

---

**CENTRAL ILLUSTRATION Early Valve Replacement vs Surveillance in Moderate Aortic Stenosis**

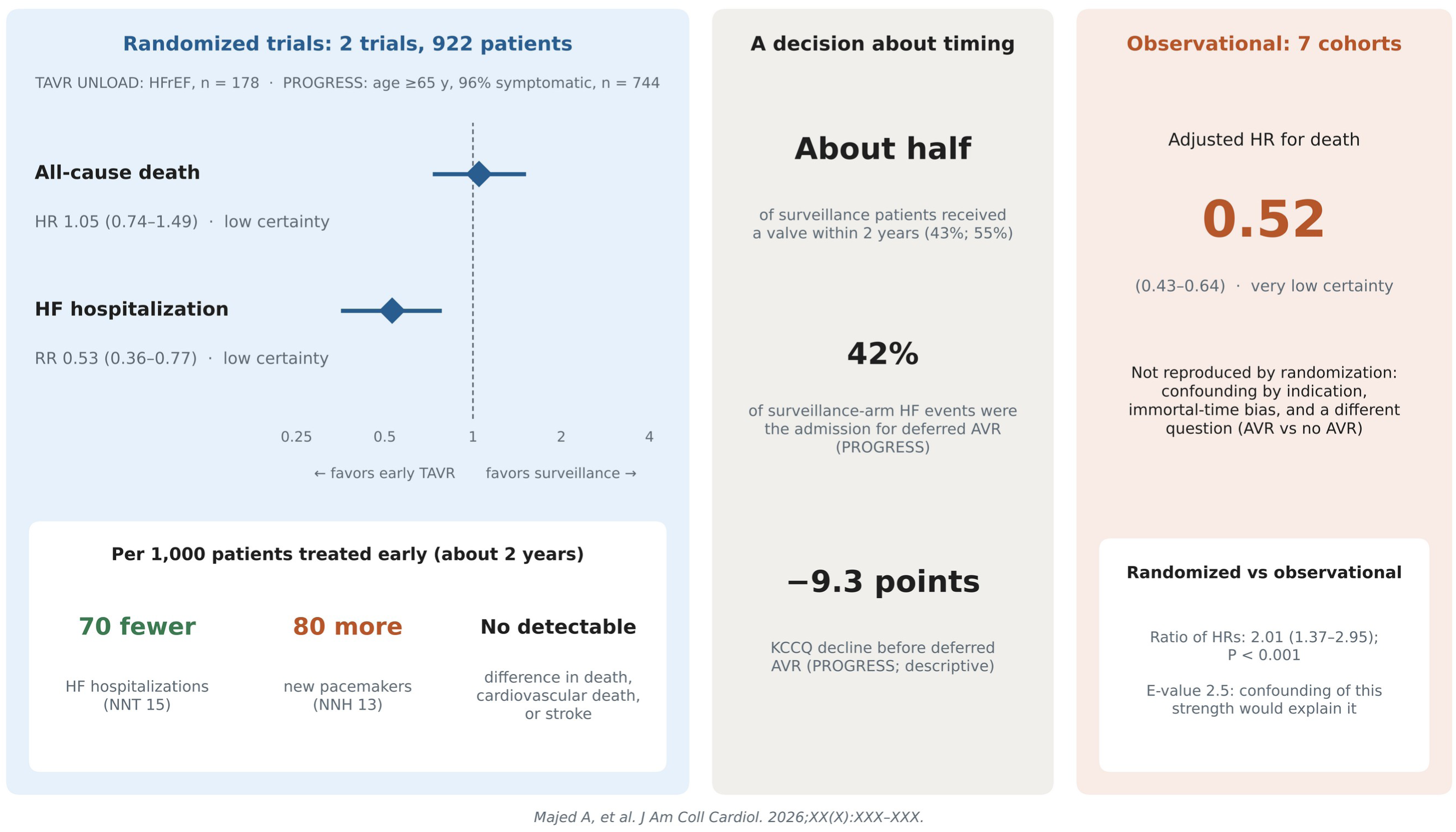

