## Supplemental Appendix for "Early Valve Replacement vs Surveillance in Moderate Aortic Stenosis: Meta-Analysis of Randomized and Observational Evidence"

### Contents

Supplemental Methods 2

Supplemental Table 1. PRISMA 2020 Checklist 5

Supplemental Table 2. Deviations From the Registered Protocol 7

Supplemental Table 3. Sources, Dates, and Yields of the Search 9

Supplemental Table 4. Outcome Definitions, How Each Trial Reported Them, and Handling in This Review 10

Supplemental Figure 1. PRISMA 2020 Flow Diagram 11

Supplemental Table 5. Reports Excluded After Full-Text Assessment, With Reasons 12

Supplemental Table 6. Design and Baseline Characteristics of the Randomized Trials 13

Supplemental Figure 2. Risk of Bias 15

Supplemental Table 7. Risk-of-Bias Judgments With Supporting Reasons 16

Supplemental Table 8. Sensitivity and Bayesian Analyses 18

Supplemental Table 9. Outcomes by Trial and Arm, and Pooled Estimates 19

Supplemental Table 10. Periprocedural (30-Day) Outcomes After Early and After Deferred Valve Replacement 20

Supplemental Table 11. Effect Modification: Between-Trial, Within-PROGRESS, and Observational Interaction Tests 21

Supplemental Table 12. Adjusted Observational Studies: Characteristics, Estimates, and Risk of Bias 22

Supplemental Table 13. GRADE Evidence Profile 23

Supplemental Table 14. Applicability of the Evidence and Ongoing Trials 25

Supplemental References 25

### Supplemental Methods

#### *Search strategies*

**PubMed (MEDLINE), November 3, 2026; 3,115 records.** ("Aortic Valve Stenosis"[Mesh] OR "aortic stenosis"[tiab] OR "aortic valve stenosis"[tiab]) AND (moderate[tiab] OR nonsevere[tiab] OR non-severe[tiab]) AND ("Transcatheter Aortic Valve Replacement"[Mesh] OR "Aortic Valve/surgery"[Mesh] OR TAVR[tiab] OR TAVI[tiab] OR "valve replacement"[tiab] OR "valve implantation"[tiab] OR AVR[tiab] OR SAVR[tiab] OR "early intervention"[tiab] OR "valve intervention"[tiab])

An initial, narrower string run on November 2, 2026 (949 records, with an additional comparator and design block) did not retrieve a relevant cohort identified from a trial reference list; the broader string above captures every record of the initial string and replaced it.

**Embase (embase.com), November 3, 2026; 3,524 records, of which 2,176 were not indexed in MEDLINE.** ('aortic valve stenosis'/exp OR 'aortic stenosis':ti,ab OR 'aortic valve stenosis':ti,ab) AND (moderate:ti,ab OR nonsevere:ti,ab OR 'non severe':ti,ab) AND ('transcatheter aortic valve implantation'/exp OR 'aortic valve replacement'/exp OR tavr:ti,ab OR tavi:ti,ab OR 'valve replacement':ti,ab OR 'valve implantation':ti,ab OR avr:ti,ab OR savr:ti,ab OR 'early intervention':ti,ab OR 'valve intervention':ti,ab) AND (surveillance:ti,ab OR conservative:ti,ab OR 'medical therapy':ti,ab OR 'medical management':ti,ab OR 'watchful waiting':ti,ab OR random*:ti,ab OR propensity:ti,ab OR matched:ti,ab OR cohort:ti,ab OR mortality:ti,ab OR survival:ti,ab)

**ClinicalTrials.gov, November 3, 2026; 15 records.** Condition "moderate aortic stenosis"; allocation randomized.

**Web of Science Core Collection, November 3, 2026; 2,433 records.** TS=("aortic stenosis" OR "aortic valve stenosis") AND TS=(moderate OR nonsevere OR "non-severe") AND TS=(TAVR OR TAVI OR "valve replacement" OR "valve implantation" OR AVR OR SAVR OR "early intervention" OR "valve intervention" OR "transcatheter aortic")

**Cochrane CENTRAL, November 3, 2026; 295 trial records.** ("aortic stenosis" OR "aortic valve stenosis"):ti,ab,kw AND (moderate OR nonsevere OR "non-severe"):ti,ab,kw AND (TAVR OR TAVI OR "valve replacement" OR "valve implantation" OR AVR OR SAVR OR "early intervention" OR "transcatheter aortic"):ti,ab,kw

**WHO International Clinical Trials Registry Platform (ICTRP) search portal, November 3, 2026; 1 record.** "moderate aortic stenosis" (the same record was among the ClinicalTrials.gov results). The portal returned only 12 records for "aortic stenosis" and did not list the trials registered at ClinicalTrials.gov, so registry coverage rests on ClinicalTrials.gov.

**Other sources.** Reference lists of the included studies and of an earlier meta-analysis of moderate aortic stenosis[S1] were screened (backward citation searching). On November 3, 2026, articles citing the 2 trials, the PROGRESS design report, the 7 included cohorts, and 2 earlier meta-analyses[S1,S2] were retrieved from Europe PMC (186 unique records) and screened by title and, where relevant, abstract (forward citation searching); 1 additional report of an included trial was identified.[S3]

#### *Keyword prefilters*

Two keyword prefilters were applied before title screening and are reported in the PRISMA flow diagram as records marked ineligible by automation tools. Embase records not indexed in MEDLINE (n = 2,176) were retained if their title or abstract mentioned moderate or non-severe stenosis together with a management comparison (143 retained). Web of Science records (n = 2,433) were retained if their title contained moderate, non-severe, pseudo-severe, or low-gradient (721 retained); of these, 284 mentioning moderate or non-severe stenosis were read in full at the title and abstract level, and abstracts or full texts were checked for 17. All 295 CENTRAL records were screened by title. Web of Science and CENTRAL records were not deduplicated against MEDLINE and Embase at the record level; neither source identified an eligible study not already found.

#### *Statistical methods*

Standard errors of log hazard ratios (HRs) were derived from published 95% CIs as (ln upper − ln lower)/(2 × 1.96). Common-effect estimates used inverse-variance weights for log HRs and the Mantel-Haenszel method for risk ratios (RRs). Between-study variance ($\tau^2$) was estimated by restricted maximum likelihood. The modified Hartung-Knapp-Sidik-Jonkman adjustment multiplied the variance of the random-effects estimate by max(1, q), where $q = \Sigma w_i(y_i - \hat{\mu})^2/(k - 1)$, and used a t distribution with $k - 1$ degrees of freedom; the unmodified version is also reported for heart failure (HF) hospitalization.

The Bayesian random-effects model used a normal likelihood for each log effect with variance $SE_i^2 + \tau^2$, a normal prior with mean 0 and SD 1 for the pooled log effect μ, and a half-normal prior for τ with scale 0.5 (0.25 and 1.0 in sensitivity analyses). The posterior was computed by integrating τ numerically (20,001 points from 0 to 12 times the prior scale) and μ analytically given τ, and was confirmed by Monte Carlo sampling. The posterior median and 2.5th and 97.5th percentiles of the marginal posterior of μ are reported, with the posterior probabilities of any reduction ($\mu < 0$) and of a reduction of at least 15% ($\mu < \ln 0.85$).

Absolute effects used the PROGRESS surveillance-arm 2-year risk $p_0$ as baseline. For HRs, the absolute difference was $1 - (1 - p_0)$^HR $- p_0$; for RRs, $p_0(RR - 1)$. Numbers needed to treat or harm are reciprocals of absolute differences, rounded up. For pacemaker implantation, the absolute effect is the difference between PROGRESS Kaplan-Meier estimates (17.3% vs 9.3%), with a CI from the binomial variance of those estimates and denominators approximated as patients without a pacemaker or implantable cardioverter-defibrillator at baseline (327 and 332).

The optimal information size for time-to-event outcomes was $4(z_{1-\alpha/2} + z_{1-\beta})^2/(\ln HR)^2$ events, with $\alpha = 0.05$, power 80%, and HR 0.80 for death (631 deaths) or 0.75 for HF events (380 events), rounded up; it was converted to patients using the average 2-year risk across arms. For HF hospitalization, the information size for a 25% relative reduction from a 14.9% baseline risk was about 2,560 patients.

Differences between 2 relative effects (between trials, between subgroups, between randomized and observational tiers) were tested as the ratio of the 2 estimates, with SE of its logarithm equal to $\sqrt{(SE_1^2 + SE_2^2)}$.[S4] Interaction P values for PROGRESS subgroups were derived from published subgroup CIs. For the observational pooled HR, the E-value was computed after converting the HR to an approximate RR as $(1 - 0.5^{\sqrt{HR})/(1 - 0.5}\sqrt{(1/HR)})$, appropriate when the outcome is common.[S5] Trial sequential analysis used Lan-DeMets O'Brien-Fleming α spending (2-sided α 0.05), with boundaries computed from the multivariate normal distribution of the cumulative Z statistics, trials ordered by publication date, and information fractions from cumulative patients (HF hospitalization) or deaths (mortality); no heterogeneity adjustment was needed ($I^2 = 0\%$).[S6] Analyses were run in Python 3 with NumPy and SciPy; the code is available at https://doi.org/10.5281/zenodo.23950111.

### Supplemental Table 1. PRISMA 2020 Checklist

| Section and item | Location in manuscript |
|---|---|
| 1 Title | Title page (identified as a meta-analysis) |
| 2 Abstract | Structured abstract |
| 3 Rationale | Introduction, paragraphs 1 and 2 |
| 4 Objectives | Introduction, paragraph 3 |
| 5 Eligibility criteria | Methods, Search and eligibility |
| 6 Information sources | Methods, Search and eligibility; Supplemental Table 3 |
| 7 Search strategy | Supplemental Methods, Search strategies |
| 8 Selection process | Methods, Selection and data extraction; Supplemental Methods, Keyword prefilters |
| 9 Data collection process | Methods, Selection and data extraction |
| 10a Data items: outcomes | Methods, Outcomes and harmonization; Supplemental Table 4 |
| 10b Data items: other variables | Table 1; Supplemental Tables 6 and 12 |
| 11 Study risk of bias assessment | Methods, Risk of bias and certainty |
| 12 Effect measures | Methods, Statistical analysis |
| 13a Synthesis eligibility | Methods, Search and eligibility; Outcomes and harmonization |
| 13b Data preparation | Methods, Outcomes and harmonization; Supplemental Methods, Statistical methods |
| 13c Tabulation and display | Tables 1 and 2; Figures 1 to 3 |
| 13d Synthesis methods | Methods, Statistical analysis; Supplemental Methods |
| 13e Heterogeneity and effect modification | Methods, Statistical analysis |
| 13f Sensitivity analyses | Methods, Statistical analysis |
| 14 Reporting bias assessment | Methods, Statistical analysis (ROB-ME) |
| 15 Certainty assessment | Methods, Risk of bias and certainty |
| 16a Study selection | Results, Study selection; Supplemental Figure 1 |
| 16b Excluded studies | Supplemental Table 5 |
| 17 Study characteristics | Table 1; Supplemental Tables 6 and 12 |
| 18 Risk of bias in studies | Results, Study selection; Supplemental Figure 2; Supplemental Table 7 |
| 19 Results of individual studies | Figures 1 to 3; Supplemental Table 9 |
| 20a Characteristics of syntheses | Results |
| 20b Results of syntheses | Results; Figures 1 and 2; Table 2 |
| 20c Heterogeneity and effect modification | Results, Effect modification; Figure 3; Supplemental Table 11 |
| 20d Sensitivity analyses | Results; Supplemental Table 8 |
| 21 Reporting biases | Results, Study selection (ROB-ME) |
| 22 Certainty of evidence | Results, Certainty of evidence; Table 2; Supplemental Table 13 |
| 23a-d Discussion | Discussion |
| 24a-b Registration and protocol | Methods, Protocol and registration (PROSPERO CRD420261559767, registered November 3, 2026) |
| 24c Amendments | Supplemental Table 2 |
| 25 Support | Funding Support and Author Disclosures |

| Section and item | Location in manuscript |
| --- | --- |
| 26 Competing interests | Funding Support and Author Disclosures |
| 27 Availability of data, code, and materials | Methods, Data sharing statement |

### Supplemental Table 2. Deviations From the Registered Protocol

| Deviation | Reason | Status |
|---|---|---|
| Trial populations classified by phenotype before synthesis | To structure interpretation of 2 trials enrolling different populations | Post hoc |
| No prediction interval reported | Not estimable with 2 trials | — |
| HF hospitalization harmonized as first disease-related event at 1 year in TAVR UNLOAD and first HF hospitalization at 2 years in PROGRESS, pooled as an RR | Only form in which TAVR UNLOAD reported hospitalization separately | Post hoc |
| Each trial's broader HF event outcome pooled as a secondary, non-harmonized outcome | Definitions differ between trials | Post hoc |
| Bayesian priors specified numerically (normal [0, 1] for the log effect; half-normal $\tau$ with scale 0.25, 0.5, 1.0) | Protocol named weakly informative priors without values | Post hoc |
| PubMed string broadened | A relevant cohort was found outside the initial string | Before extraction |
| Keyword prefilters applied to Embase-only and Web of Science records | Volume of records; recorded as automation in the PRISMA flow diagram | Before screening |
| Observational conference abstracts excluded from synthesis and summarized narratively; the one abstract with an adjusted estimate added in a sensitivity analysis | Not peer reviewed; limited methods; possible overlap with published cohorts | Post hoc |
| Ratio of HRs between tiers, E-values, optimal information size and trial sequential analysis, ICEMAN credibility assessment, and inclusion of an excluded cohort as sensitivity analysis | Requested in internal review | Post hoc |
| Planned reconstruction of individual participant data from Kaplan-Meier curves (one-stage Cox model stratified by trial; restricted mean survival time at 1 and 2 y) not performed; review title changed accordingly | Pooled trial-level estimates and a common 2-year horizon address the same question with less digitization error | — |
| Outcomes at 1 y (registered alongside longest follow-up) not analyzed; death also analyzed at a common 2-y horizon | PROGRESS reported clinical outcomes at 2 y; 1-y estimates would have required digitizing Kaplan-Meier curves | Post hoc |
| Registered subgroups by symptom status and by transcatheter vs surgical AVR not analyzable; sex, NYHA class, atrial fibrillation, valve area, peak velocity, NT-proBNP, calcium score, surgical risk, and cardiac damage taken from PROGRESS's published subgroup analysis | 96% of PROGRESS patients were symptomatic, and all early AVR in both trials was transcatheter | Post hoc |
| KCCQ change not pooled as a mean difference | Measured at different time points (1 y and 2 y) | — |
| Trial investigators not contacted for unreported data (eg, HF hospitalization at 2 y in TAVR UNLOAD); late-breaking congress sessions not hand-searched | Not done before submission; congress abstracts were captured through Embase | — |
| WHO ICTRP searched through its portal with limited yield (Supplemental Methods) | Portal returned incomplete results | — |
| Analyses run in Python (NumPy, SciPy, statsmodels, PyMARE) rather than R | Scripts reproduce the registered models and are publicly available | — |

AVR = aortic valve replacement; HF = heart failure; HR = hazard ratio; ICEMAN = Instrument for assessing the Credibility of Effect Modification Analyses; KCCQ = Kansas City Cardiomyopathy Questionnaire; NT-proBNP = N-terminal pro-B-type natriuretic peptide;

NYHA = New York Heart Association; RR = risk ratio; WHO ICTRP = World Health Organization International Clinical Trials Registry Platform.

### Supplemental Table 3. Sources, Dates, and Yields of the Search

| Source | Date | Concept blocks | Records | Handling before screening |
|---|---|---|---|---|
| PubMed (MEDLINE) | November 3, 2026 | Stenosis AND moderate AND valve replacement | 3,115 | All screened |
| Embase | November 3, 2026 | As above, plus comparator or design block | 3,524 | 1,348 indexed in MEDLINE removed; 2,176 Embase-only prefiltered to 143 |
| ClinicalTrials.gov | November 3, 2026 | Moderate aortic stenosis; randomized | 15 | All screened |
| WHO ICTRP search portal | November 3, 2026 | Moderate aortic stenosis | 1 | Duplicate of a ClinicalTrials.gov record; removed |
| Web of Science Core Collection | November 3, 2026 | Stenosis AND moderate AND valve replacement | 2,433 | Prefiltered by title to 721 |
| Cochrane CENTRAL | November 3, 2026 | Stenosis AND moderate AND valve replacement | 295 | All screened by title |
| Citation searching, backward | November 2026 | Reference lists of included studies and an earlier meta-analysis | 4 reports | Assessed in full |
| Citation searching, forward (Europe PMC) | November 3, 2026 | Articles citing the trials, the PROGRESS design report, the 7 cohorts, and 2 earlier meta-analyses | 186 records | Screened by title or abstract; 1 report assessed in full |

### Supplemental Table 4. Outcome Definitions, How Each Trial Reported Them, and Handling in This Review

| Outcome | TAVR UNLOAD | PROGRESS | Handling in this review |
|---|---|---|---|
| All-cause death | Time to event, longest follow-up (median 23 mo); per-patient listing with day of death | Time to event, 2 y | Pooled as HR (primary); RR at a common 2-y horizon (sensitivity) |
| Cardiovascular death | Classified per patient in the listing | Counts at 2 y; unknown causes classified as cardiovascular | Pooled as RR at longest follow-up and at 2 y |
| HF hospitalization | First disease-related event that was an HF admission, 1 y (supplementary table); later admissions after a different first event not counted | Time to first HF hospitalization, 2 y; recurrent admissions by joint frailty model | Pooled as RR (primary, harmonized); PROGRESS-only sensitivity analysis |
| HF event (trial-defined) | Disease-related hospitalization or HF equivalent (urgent visit, outpatient intensification of HF therapy) | HF event with or without hospitalization (HF Collaboratory-Academic Research Consortium; deferred AVR alone did not qualify)[S7] | Pooled as HR (secondary, non-harmonized) |
| Stroke | Any stroke, longest follow-up | Any and disabling stroke, 2 y (VARC-3)[S8] | Any stroke pooled as RR |
| New permanent pacemaker | 30 d after index TAVR only | 2 y, intention to treat, patients without a baseline pacemaker; also 30 d after early and deferred AVR | PROGRESS only |
| Bleeding, myocardial infarction, atrial fibrillation, endocarditis | 30 d after index TAVR only | 2 y, intention to treat | PROGRESS only |
| Primary composite | Hierarchical (death, disabling stroke, disease-related hospitalization or equivalent, KCCQ change); win ratio[S9] | Death or HF event, time to first event, 2 y | Reported as published; not pooled |
| Health status | KCCQ overall summary score, change at 1 y | KCCQ overall summary score, change at 2 y; alive with sustained ≥10-point gain | Reported per trial; not pooled (different time points) |
| AVR in surveillance arm | Proportion, timing, indication | Proportion, timing, indication | Reported per trial |

AVR = aortic valve replacement; HF = heart failure; HR = hazard ratio; KCCQ = Kansas City Cardiomyopathy Questionnaire; RR = risk ratio; TAVR = transcatheter aortic valve replacement; VARC-3 = Valve Academic Research Consortium 3.

### Supplemental Figure 1. PRISMA 2020 Flow Diagram

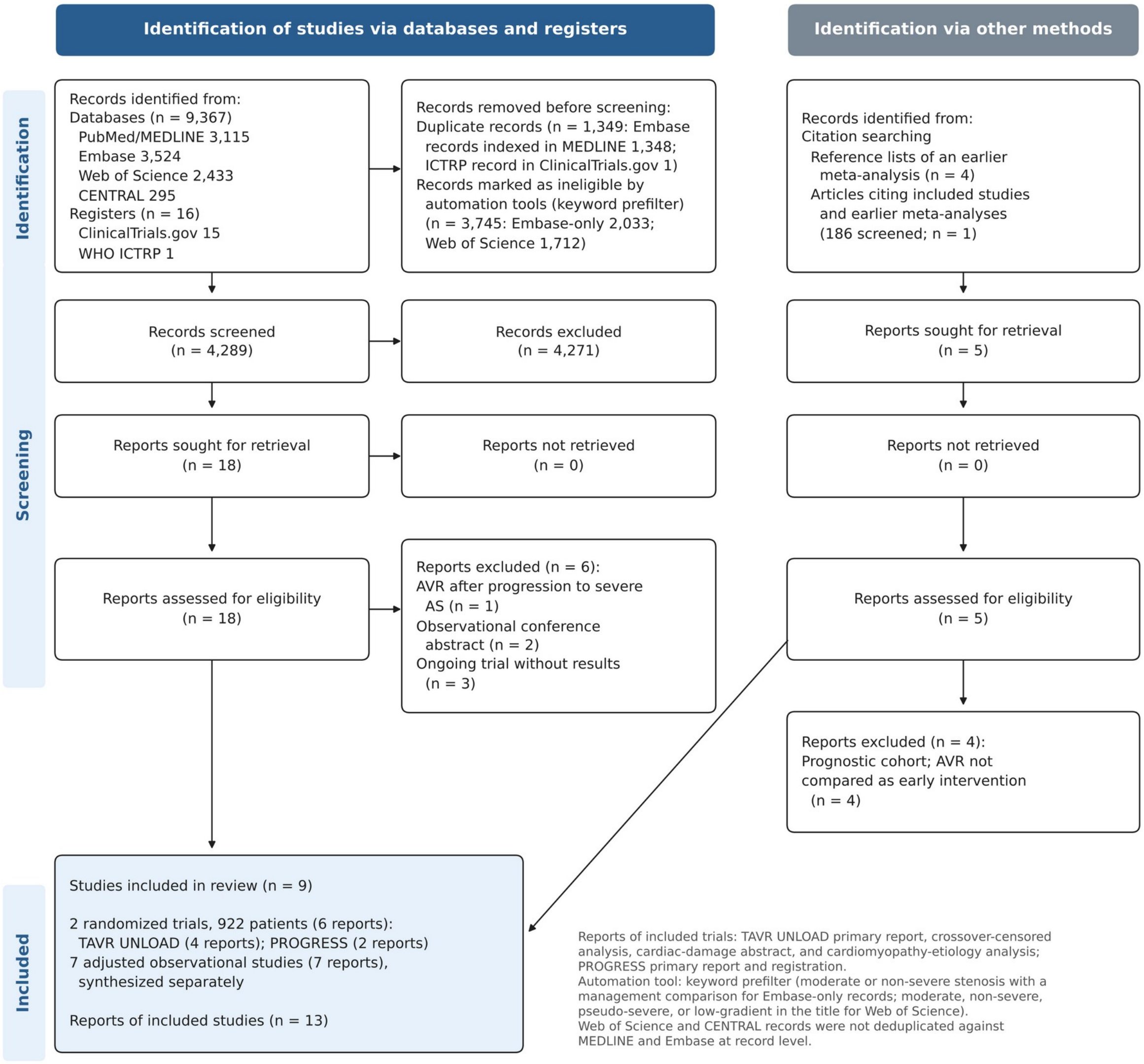


Counts reflect consensus between the 2 independent reviewers.

### Supplemental Table 5. Reports Excluded After Full-Text Assessment, With Reasons

| Report | Source | Reason for exclusion |
|---|---|---|
| Jean et al, 2021[S10] | Databases | AVR performed after progression to severe aortic stenosis and modeled as a time-dependent exposure; does not test early AVR (used in a sensitivity analysis) |
| Sabri et al, 2025 (TCT abstract)[S11] | Databases | Observational conference abstract (TriNetX; propensity-matched relative risk of death 0.79 [0.73-0.86]); not peer reviewed, risk ratio rather than HR, possible overlap with published cohorts; summarized narratively and added in a sensitivity analysis (Supplemental Table 8) |
| TOPAS registry, 2019 (ESC Congress abstract) | Databases | Observational conference abstract; pseudo-severe aortic stenosis |
| EXPAND TAVR II (NCT05149755) | Registers | Ongoing randomized trial; self-expanding valve; no results |
| TIAMAR (NCT05310461) | Registers | Ongoing randomized trial of early vs deferred AVR in moderate aortic stenosis with mitral regurgitation; no results |
| NCT07532317 | Registers | Planned randomized trial of TAVR (Myval) vs guideline-directed therapy; not yet recruiting |
| Delesalle et al, 2019 | Citation searching[S1] | Prognostic cohort; AVR modeled as a time-dependent covariate, not compared as early intervention |
| Amanullah et al, 2021 | Citation searching[S1] | As above |
| Chew et al, 2021 | Citation searching[S1] | As above |
| Du et al, 2021 | Citation searching[S1] | As above |

AVR = aortic valve replacement; TAVR = transcatheter aortic valve replacement.

### Supplemental Table 6. Design and Baseline Characteristics of the Randomized Trials

| Characteristic | TAVR UNLOAD | PROGRESS |
|---|---|---|
| Publication | J Am Coll Cardiol 2025[S12] | N Engl J Med 2026[S13] |
| Design; sponsor | Open label; investigator-initiated | Open label; valve manufacturer |
| Sites; enrollment | 66 screening institutions in the United States, the Netherlands, and Austria; 2017-2022 | 74 sites in 6 countries (87.1% United States); 2021-2024 |
| Phenotype | HFrEF on guideline-directed therapy, LVEF 20%-50%, NYHA II-IV | Age ≥65 y and ≥1 at-risk feature: symptoms, LVEF <60%, diastolic dysfunction ≥ grade 2, SVi <35 mL/m², atrial fibrillation, NT-proBNP >3× ULN, or high calcium score |
| Definition of moderate AS | AVA >1.0 and ≤1.5 cm² at rest; DSE when low flow was suspected (performed in 44.9%); indexed-AVA criteria; core laboratory | Echocardiographic moderate AS, core laboratory; 91.6% concordant on area and velocity or gradient |
| Planned vs randomized sample size | 600 planned, revised to 302; 178 randomized when enrollment ended | 628 planned; 744 randomized |
| Valve; access | Balloon-expandable; transfemoral | Balloon-expandable; transfemoral |
| n (TAVR/surveillance) | 89/89 | 373/371 |
| Age, y | 76.6 vs 78.2 | 78.5 vs 78.3 |
| Women | 18.0% vs 23.6% | 38.1% vs 40.2% |
| STS-PROM, % | 4.0 vs 4.8 | 3.1 vs 3.0 |
| NYHA II-IV; III-IV | 98.9%; 55.6% | 96.4%; 35.5% |
| HF hospitalization in prior 12 mo | 44.9% | Not reported |
| Atrial fibrillation (history) | 50.8% | 34.0% vs 32.1% |
| Coronary artery disease | 76.4% | Prior myocardial infarction 14.4%; prior PCI 30.6%; prior CABG 14.2% |
| LVEF | Mean 39.0% | 63.2% vs 63.6%; <50% in 8.5%; ≤40% in 3.0% |
| AVA; mean gradient | 1.2 cm²; 18.8 mm Hg at rest (21.9 at rest or DSE) | 1.2 cm²; 25.1 vs 25.4 mm Hg; peak velocity 3.3 m/s |
| NT-proBNP, median | Not reported | 349 vs 364 pg/mL |
| Other markers | Implantable cardioverter-defibrillator 34.8%; frailty 26.9% | Diastolic dysfunction ≥ grade 2 44.5% vs 43.1%; ≥3 at-risk features 41.0% vs 38.8%; bicuspid valve 8.0% vs 6.7% |
| KCCQ at baseline | 55.8 | 64.5 vs 63.9 |
| Follow-up | Median 23 mo | 2 y |
| AVR in surveillance arm | 38 (42.7%), median 12 mo; 35 for progression to severe AS | 204 (55.0%; Kaplan-Meier 57.4% at 2 y), 99 within 1 y; 166 for progression with a class 1 indication; 199 transcatheter |
| Events committee | Independent; blinding not stated | Independent; aware of assignment |

Values are TAVR vs surveillance where 2 values are given and both arms combined otherwise. AS = aortic stenosis; AVA = aortic valve area; CABG = coronary artery bypass grafting; DSE = dobutamine stress echocardiography; HF = heart failure; HFrEF = heart failure with reduced ejection fraction; KCCQ = Kansas City Cardiomyopathy Questionnaire; LVEF = left ventricular ejection fraction; NT-proBNP = N-terminal pro-B-type natriuretic peptide; NYHA = New York

Heart Association; PCI = percutaneous coronary intervention; STS-PROM = Society of Thoracic Surgeons Predicted Risk of Mortality; SVi = stroke volume index; TAVR = transcatheter aortic valve replacement; ULN = upper limit of normal.

**Supplemental Figure 2. Risk of Bias**

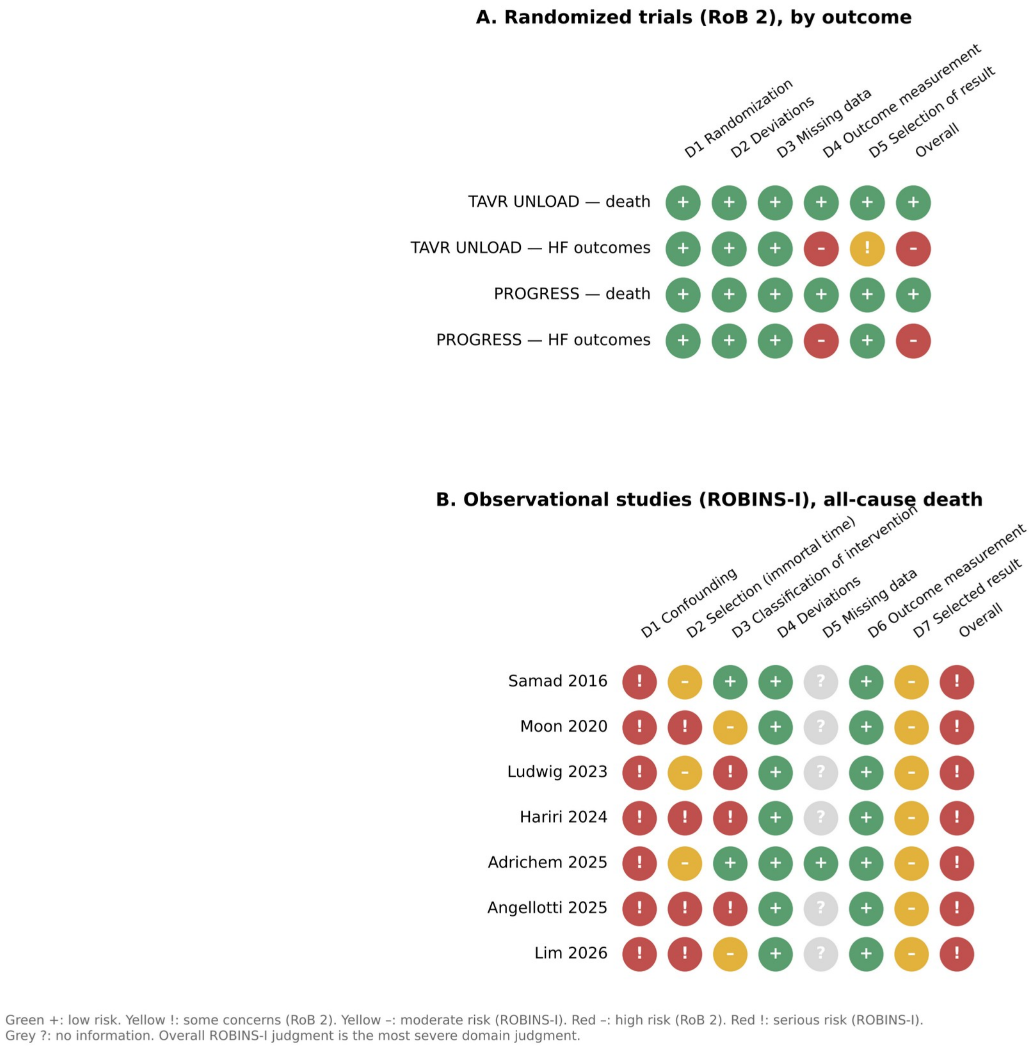


(A) RoB 2 for the randomized trials, by outcome. (B) ROBINS-I for the observational studies (all-cause death). Judgments with supporting reasons are given in Supplemental Table 7. Judgments are consensus judgments of the 2 independent reviewers.

### Supplemental Table 7. Risk-of-Bias Judgments With Supporting Reasons

| Study (outcome) | D1 | D2 | D3 | D4 | D5 | D6 | D7 | Overall | Support |
|---|---|---|---|---|---|---|---|---|---|
| TAVR UNLOAD (death) | Low | Low | Low | Low | Low | — | — | Low | Central randomization; crossover on progression permitted by protocol and treated as part of the surveillance strategy; follow-up >90%; death objective |
| TAVR UNLOAD (HF outcomes) | Low | Low | Low | High | Some concerns | — | — | High | Open label; blinding of event adjudication not stated; composite includes urgent visits and medication intensification; sample size and timing of primary analysis changed during enrollment |
| PROGRESS (death) | Low | Low | Low | Low | Low | — | — | Low | Central randomization; prespecified analysis; death objective |
| PROGRESS (HF outcomes) | Low | Low | Low | High | Low | — | — | High | Open label; events committee aware of assignment; HF event definition depends on symptoms and treatment decisions |
| Samad 2016 (death) | Serious | Moderate | Low | Low | No information | Low | Moderate | Serious | Confounding by indication; AVR within 90 days with inverse probability weighting, follow-up from AVR or day 15 (short window of misaligned time zero); concomitant CABG |
| Moon 2020 (death) | Serious | Serious | Moderate | Low | No information | Low | Moderate | Serious | AVR group 8 years younger; time zero at index echocardiogram with surgery up to 2 years later (median 20 days; immortal time) |
| Ludwig 2023 (death) | Serious | Moderate | Serious | Low | No information | Low | Moderate | Serious | Treated patients had pseudo-severe low-gradient AS managed as severe; comparator moderate AS; confounding |
| Hariri 2024 (death) | Serious | Serious | Serious | Low | No information | Low | Moderate | Serious | Surveillance defined by never receiving AVR during follow-up; follow-up began at AVR in the AVR group but at the index echocardiogram in the surveillance group (misaligned time zero); 26% with other cardiac surgery |
| Adrichem 2025 (death) | Serious | Moderate | Low | Low | Low | Low | Moderate | Serious | Crude HR 0.92 became 0.51 after adjustment for an older, far more symptomatic treated group; vital status from the municipal registry; residual confounding |
| Angellotti 2025 (death) | Serious | Serious | Serious | Low | No information | Low | Moderate | Serious | Any AVR during follow-up (median 749 days), not time-dependent; 19% no longer moderate at AVR; adjustment limited to 5 variables |
| Lim 2026 (death) | Serious | Serious | Moderate | Low | No information | Low | Moderate | Serious | AVR within 1 year (median 22 days) of index echocardiogram with time zero at the echocardiogram (immortal time); confounding |

RoB 2 domains for trials: D1 randomization process; D2 deviations from intended interventions (effect of assignment); D3 missing outcome data; D4 measurement of the outcome; D5 selection of the reported result. ROBINS-I domains for observational studies: D1 confounding; D2 selection of participants (including immortal time); D3 classification of interventions; D4 deviations from intended interventions; D5 missing data; D6 measurement of outcomes; D7 selection of the reported result. ROBINS-I D4 was judged for the effect of starting AVR, so later AVR in comparator patients was regarded as usual practice; D6 was low because all-cause death is objective; D7 was moderate because no prespecified analysis plan was available. D5 was judged no information where a cohort did not report missing data. Judgments are consensus judgments of the 2 independent reviewers. AS = aortic stenosis; AVR = aortic valve replacement; CABG = coronary artery bypass grafting; HF = heart failure; HR = hazard ratio.

### Supplemental Table 8. Sensitivity and Bayesian Analyses

| Analysis | Estimate (95% CI or credible interval) |
|---|---|
| Death, common effect (primary) | HR 1.05 (0.74-1.49) |
| Death, random effects (REML, modified HKSJ) | HR 1.05 (0.11-10.20) |
| Death, Mantel-Haenszel RR, longest follow-up | RR 1.06 (0.79-1.44) |
| Death, common 2-y horizon | RR 1.19 (0.83-1.71) |
| Death, Bayesian (τ scale 0.25 / 0.5 / 1.0) | HR 1.05 (0.67-1.64) / 1.05 (0.58-1.88) / 1.05 (0.47-2.28); P(any reduction) 41% / 42% / 44%; P(≥15% reduction) 16% / 20% / 24% |
| Death, absolute effect per 1,000 at 2 y: PROGRESS-like / TAVR UNLOAD-like / pooled crude 2-y risk | 3 more (19 fewer to 34 more) / 9 more (54 fewer to 91 more) / 4 more (25 fewer to 45 more) |
| HF hospitalization, Mantel-Haenszel (primary) | RR 0.53 (0.36-0.77) |
| HF hospitalization, inverse variance | RR 0.53 (0.36-0.78) |
| HF hospitalization, random effects (REML, modified HKSJ / unmodified HKSJ) | RR 0.53 (0.04-6.32) / 0.53 (0.15-1.93) |
| HF hospitalization, Bayesian (τ scale 0.25 / 0.5 / 1.0) | RR 0.55 (0.35-0.92) / 0.56 (0.32-1.15) / 0.58 (0.28-1.58); P(any reduction) 99% / 95% / 90% |
| HF hospitalization, PROGRESS only | RR 0.50 (0.32-0.78); recurrent events HR 0.52 (0.35-0.78) |
| HF events (trial-defined), random effects (REML, modified HKSJ) | HR 0.65 (0.06-6.95) |
| HF events, Bayesian (τ scale 0.5) | HR 0.67 (0.38-1.30); P(any reduction) 92% |
| Cardiovascular death, common 2-y horizon | RR 1.00 (0.63-1.58) |
| Observational, leave-one-out range | HR 0.49-0.55 |
| Observational, excluding time-related bias (Moon, Hariri, Angellotti, Lim) | HR 0.50 (0.29-0.86) |
| Observational, alternative adjusted estimates (Samad 0.68; Moon 0.70) | HR 0.56 (0.45-0.69) |
| Observational, excluding concomitant surgery (Samad, Hariri, Angellotti) | HR 0.51 (0.32-0.80) |
| Observational, excluding pseudo-severe AS (Ludwig) | HR 0.55 (0.46-0.68) |
| Observational, including Jean 2021 | HR 0.53 (0.44-0.63) |
| Observational, including the Sabri 2025 abstract (risk ratio treated as HR) | HR 0.56 (0.44-0.71); $I^2 = 77\%$; ratio of HRs vs randomized 1.87 (1.25-2.79); $P = 0.002$ |
| Trial sequential analysis, death (information size 631 deaths; 20% of it reached) | Cumulative Z 0.26; O'Brien-Fleming boundary 4.39 at this information fraction; not crossed |
| Trial sequential analysis, HF hospitalization (information size about 2,560 patients; 36% reached) | Cumulative Z 3.28; boundary 3.26; marginally crossed |
| Randomized vs observational, ratio of HRs for death | 2.01 (1.37-2.95); $P < 0.001$ |
| E-value for the observational HR (point estimate / CI bound) | 2.5 / 2.1 |

AS = aortic stenosis; HF = heart failure; HKSJ = Hartung-Knapp-Sidik-Jonkman; HR = hazard ratio; REML = restricted maximum likelihood; RR = risk ratio.

### Supplemental Table 9. Outcomes by Trial and Arm, and Pooled Estimates

| Outcome | TAVR UNLOAD, TAVR vs surveillance | PROGRESS, TAVR vs surveillance | Pooled (95% CI) |
|---|---|---|---|
| All-cause death, longest follow-up | 35/89 vs 35/89; HR 0.98 (0.61-1.56) | 30/373 vs 26/371; HR 1.14 (0.67-1.93) | HR 1.05 (0.74-1.49) |
| All-cause death, 2-y horizon | 25/89 vs 20/89* | 30/373 vs 26/371 | RR 1.19 (0.83-1.71) |
| Cardiovascular death, longest follow-up | 28/89 vs 29/89* | 11/373 vs 14/371 | RR 0.91 (0.62-1.32) |
| Cardiovascular death, 2-y horizon | 20/89 vs 17/89* | 11/373 vs 14/371 | RR 1.00 (0.63-1.58) |
| Non-cardiovascular death, 2-y horizon | 5/89 vs 3/89* | 19/373 vs 12/371 | RR 1.59 (0.85-3.00) |
| HF hospitalization | 10/89 vs 16/89 (first event, 1 y) | 26/373 vs 52/371 (2 y) | RR 0.53 (0.36-0.77) |
| HF hospitalizations per 100 patient-years | — | 6.2 vs 12.0; HR 0.52 (0.35-0.78) | — |
| HF event, trial-defined | 34/89 vs 40/89; HR 0.80 (0.51-1.27) | 38/373 vs 66/371; HR 0.55 (0.37-0.81) | HR 0.64 (0.48-0.87) |
| Any stroke | 2/89 vs 6/89 | 20/373 vs 16/371 | RR 1.00 (0.56-1.77) |
| Disabling stroke, 2 y | — | 10/373 vs 7/371 | — |
| Myocardial infarction, 2 y | — | 7/373 vs 11/371 | — |
| Life-threatening or fatal bleeding, 2 y | — | 17/373 vs 18/371 | — |
| New atrial fibrillation, 2 y | — | 28/246 vs 22/252 (11.7% vs 9.3%)† | — |
| New permanent pacemaker, 2 y | — | 55 vs 29 (17.3% vs 9.3%)‡ | — |
| Endocarditis, 2 y | — | 4/373 vs 3/371 | — |
| Primary endpoint as defined by trial | Win ratio 1.31 (0.91-1.88) | Death or HF event: 16.7% vs 23.9%; HR 0.67 (0.48-0.93) | Not pooled |
| Death, stroke, or HF hospitalization | — | 17.7% vs 23.3%; P = 0.07 | — |
| Health status | KCCQ +12.8 vs +3.2 at 1 y; difference 9.6 (1.7-17.4) | KCCQ +14.3 vs +7.7 at 2 y; sustained ≥10-point gain 134/344 vs 76/327 | Not pooled |
| NYHA III-IV at 2 y | — | 3.9% vs 10.9% | — |
| AVR in surveillance arm | 38/89 (42.7%) | 204/371 (55.0%; Kaplan-Meier 57.4%) | 242/460 (52.6%) |

*Counted from the trial's per-patient listing of deaths. †Patients without atrial fibrillation at baseline; percentages are Kaplan-Meier estimates. ‡Patients without a pacemaker at baseline; Kaplan-Meier estimates. Pooled death HR by inverse variance; RRs by Mantel-Haenszel. AVR = aortic valve replacement; HF = heart failure; HR = hazard ratio; KCCQ = Kansas City Cardiomyopathy Questionnaire; NYHA = New York Heart Association; RR = risk ratio; TAVR = transcatheter aortic valve replacement.

**Supplemental Table 10. Periprocedural (30-Day) Outcomes After Early and After Deferred Valve Replacement**

| Outcome | TAVR UNLOAD, early TAVR (n = 88) | PROGRESS, early TAVR (n = 358) | PROGRESS, deferred AVR within 2 y (n = 204) |
|---|---|---|---|
| Death | 0 | 0 | 1 (0.5%) |
| Stroke (disabling) | 1 (1.1%) (1) | 3 (0.8%) (0) | 5 (2.5%) (3) |
| Major bleeding | — | 1 (0.3%) | 5 (2.5%) |
| Life-threatening or fatal bleeding | 1 (1.1%)* | 2 (0.6%) | 5 (2.5%) |
| Vascular complication, any (major) | Not reported (0) | 7 (2.0%) (2) | 14 (6.9%) (3) |
| Acute kidney injury | 1 (1.1%)† | 3 (0.8%) | 3 (1.5%) |
| New permanent pacemaker | 4 (4.5%)‡ | 41 (13.0%)§ | 19 (10.4%)§ |
| New atrial fibrillation§ | 0 | 11 (4.6%) | 8 (6.1%) |
| Myocardial infarction | — | 1 (0.3%) | 2 (1.0%) |
| Reintervention for valve dysfunction | 1 (1.1%) | — | — |

Descriptive, nonrandomized comparison: deferred procedures were triggered by progression and were sometimes urgent. Percentages for PROGRESS are Kaplan-Meier estimates. *Bleeding Academic Research Consortium type 3-5. †Stage 2 or 3. ‡New pacemaker or cardiac resynchronization device among all treated patients. §Among patients without the condition at baseline (for deferred AVR, at the preprocedure visit). AVR = aortic valve replacement; TAVR = transcatheter aortic valve replacement.

### Supplemental Table 11. Effect Modification: Between-Trial, Within-PROGRESS, and Observational Interaction Tests

| Comparison | Estimates | Ratio (95% CI) | P |
|---|---|---|---|
| Between trials: all-cause death | HR 0.98 (TAVR UNLOAD) vs 1.14 (PROGRESS) | 0.86 (0.42-1.74) | 0.68 |
| Between trials: HF hospitalization | RR 0.63 vs 0.50 | 1.26 (0.53-2.97) | 0.60 |
| Between trials: trial-defined HF events | HR 0.80 vs 0.55 | 1.45 (0.80-2.65) | 0.22 |
| PROGRESS: age <75 vs ≥75 y | HR 0.52 (0.22-1.24) vs 0.70 (0.49-1.00) | 1.35 | 0.53 |
| PROGRESS: women vs men | HR 0.75 (0.45-1.26) vs 0.62 (0.40-0.95) | 0.83 | 0.58 |
| PROGRESS: surgical risk low / intermediate / high | HR 0.41 (0.21-0.81) / 0.86 (0.51-1.45) / 0.58 (0.33-1.03) | — | 0.22* |
| PROGRESS: NYHA I-II vs III-IV | HR 0.70 (0.45-1.08) vs 0.63 (0.38-1.05) | 0.90 | 0.76 |
| PROGRESS: no atrial fibrillation vs atrial fibrillation | HR 0.56 (0.36-0.87) vs 0.83 (0.50-1.38) | 1.48 | 0.25 |
| PROGRESS: AVA ≤1.2 vs >1.2 cm² | HR 0.66 (0.43-1.01) vs 0.67 (0.39-1.15) | 1.02 | 0.97 |
| PROGRESS: peak velocity <3 vs ≥3 m/s | HR 0.73 (0.36-1.50) vs 0.64 (0.44-0.93) | 0.88 | 0.75 |
| PROGRESS: LVEF <50% vs ≥50% | HR 0.71 (0.31-1.63) vs 0.65 (0.46-0.93) | 0.92 | 0.85 |
| PROGRESS: cardiac damage stage 0-1 vs 2-4 | HR 0.22 (0.06-0.79) vs 0.75 (0.52-1.08); 16 vs 113 events | 3.41 (0.89-13.0) | 0.07 |
| PROGRESS: NT-proBNP <3× vs ≥3× ULN | HR 0.68 (0.44-1.05) vs 0.66 (0.40-1.10) | 0.97 | 0.93 |
| PROGRESS: calcium score not elevated vs elevated | HR 0.66 (0.45-0.96) vs 0.70 (0.36-1.36) | 1.06 | 0.88 |
| Lim 2026: damage stage 0-2 vs 3-4 (death) | HR 0.76 (0.52-1.09) vs 0.45 (0.26-0.79) | 0.59 (0.30-1.15) | 0.12 |
| Angellotti 2025: early vs advanced damage (death) | HR 0.61 (0.25-1.44) vs 0.43 (0.28-0.66) | 0.70 (0.27-1.87) | 0.48 |
| TAVR UNLOAD: left-heart (stage 1-2) vs right-heart (stage 3-4) damage[S14] | Major adverse events at 1 y; KCCQ change (greater benefit with right-heart damage) | — | 0.27; 0.01 |
| TAVR UNLOAD: ischemic vs nonischemic cardiomyopathy[S3] | 1-y composite clinical endpoint; KCCQ and LVEF change | — | 0.85; not significant |

PROGRESS subgroup HRs are for the trial's primary endpoint (death or HF event), read from its published forest plot; ratios are the second subgroup relative to the first, and P values were derived from the published CIs. *Heterogeneity across 3 groups ($\chi^2$, 2 df). ICEMAN assessment of effect modification by cardiac damage stage: within-trial comparison (yes); direction hypothesized a priori (no; prior observational evidence suggested the opposite); supported by prior evidence (no); chance unlikely by the interaction test (no; P = 0.07); small number of effect modifiers tested (no; 11 variables); arbitrary cut points avoided (stages defined by the trial). Overall credibility: low.[S15] AVA = aortic valve area; HF = heart failure; HR = hazard ratio; KCCQ = Kansas City Cardiomyopathy Questionnaire; LVEF = left ventricular ejection fraction; NT-proBNP = N-terminal pro-B-type natriuretic peptide; NYHA = New York Heart Association; RR = risk ratio; ULN = upper limit of normal.

### Supplemental Table 12. Adjusted Observational Studies: Characteristics, Estimates, and Risk of Bias

| Study | Population; n (AVR/surveillance) | Definition of early AVR; procedure | Analysis | Adjusted HR for death (95% CI) | Principal bias concerns | ROBINS-I overall |
|---|---|---|---|---|---|---|
| Samad 2016[S16] | Moderate AS (mean gradient), LVEF ≤50%; single-center database; n = 1,090 | AVR within 90 days; SAVR with or without CABG | Cox with inverse probability of treatment weighting; time zero at AVR (recipients) or day 15 (non-recipients) | 0.59 (0.44-0.78); time-dependent model for AVR within 5 y: 0.68 (0.52-0.90) | Concomitant CABG; confounding | Serious |
| Moon 2020[S17] | AVA 1.0-1.5 cm², LVEF <50%; 37/218 | SAVR within 2 y of index echocardiogram (median 20 days) | Multivariable Cox (primary); time zero at index echocardiogram | 0.43 (0.20-0.91); with inverse probability weighting: 0.70 (0.49-0.99) | Immortal time; AVR group 8 y younger | Serious |
| Ludwig 2023[S18] | LVEF <50%; pseudo-severe low-gradient AS treated vs medically managed moderate AS | TAVR (managed as severe) | Multivariable Cox in all nonsevere AS (primary); propensity matching | 0.39 (0.27-0.55); matched: 0.38 (0.24-0.61) | Treated population classified as severe; confounding | Serious |
| Hariri 2024[S19] | Moderate AS, all LVEF; 299/1,122 | Any AVR during follow-up while still moderate (73.6% SAVR; 26% with other cardiac surgery) | Multivariable Cox; follow-up from AVR (AVR group) or index echocardiogram (surveillance); propensity matching as sensitivity | 0.51 (0.34-0.77) | Ever-treated classification; misaligned time zero; concomitant surgery | Serious |
| Adrichem 2025[S20] | Moderate AS (peak velocity 2.5-4 m/s, AVA >1.0 cm²), all LVEF; 115/220 | TAVR at moderate stage | Multivariable Cox (primary); propensity matching | 0.51 (0.35-0.76); crude 0.92 (0.68-1.26); matched, NYHA-adjusted: 0.57 (0.35-0.91) | Treated group older and far more symptomatic | Serious |
| Angellotti 2025[S21] | Moderate AS (AVA ≤1.5 cm² or mean gradient ≥20 mm Hg), all LVEF; 2 registries; 122/221 | Any AVR during follow-up (median 749 days; 81% still moderate at AVR); 70% TAVR; 8% with CABG | Cox adjusted for age, sex, gradient, SVi, and registry; AVR not time-dependent | 0.46 (0.31-0.68), strata combined; early damage 0.61 (0.25-1.44); advanced 0.43 (0.28-0.66) | Immortal time; minimal adjustment | Serious |
| Lim 2026[S22] | Moderate AS, all LVEF; 2,490 (211 early) | Any AVR within 1 y of index echocardiogram (median 22 days) | Multivariable Cox; propensity matching | 0.67 (0.50-0.92); stage 3-4: 0.45 (0.26-0.79); stage 0-2: 0.76 (0.52-1.09) | Immortal time; confounding | Serious |
| Jean 2021[S10] (excluded; sensitivity analysis only) | Moderate AS, LVEF <50% | AVR after progression to severe AS, time-dependent | Time-dependent Cox | 0.59 (0.35-0.98) | Does not test early AVR | — |

Pooled random-effects HR for the 7 included studies: 0.52 (0.43-0.64); $I^2$ = 8%. AS = aortic stenosis; AVA = aortic valve area; AVR = aortic valve replacement; CABG = coronary artery bypass grafting; HR = hazard ratio; LVEF = left ventricular ejection fraction; SAVR = surgical aortic valve replacement; SVi = stroke volume index; TAVR = transcatheter aortic valve replacement.

### Supplemental Table 13. GRADE Evidence Profile

| Outcome | Studies (design) | Risk of bias | Inconsistency | Indirectness | Imprecision | Other | Relative effect (95% CI) | Absolute effect per 1,000 at 2 y | Certainty |
|---|---|---|---|---|---|---|---|---|---|
| All-cause death | 2 (RCT) | Not serious | Not serious ($I^2$ = 0%) | Not serious* | Very serious: CI includes 26% reduction and 49% increase; 126 of 631 deaths needed | None | HR 1.05 (0.74-1.49) | 3 more (19 fewer to 34 more) | Low |
| HF hospitalization | 2 (RCT) | Serious: open label; unblinded adjudication | Not serious ($I^2$ = 0%) | Not serious† | Serious: 922 of about 2,560 patients needed; random-effects intervals include no effect | Outside PROGRESS testing hierarchy | RR 0.53 (0.36-0.77) | 70 fewer (95 to 34 fewer) | Low |
| HF event, trial-defined | 2 (RCT) | Serious | Not serious ($I^2$ = 33%) | Serious: definitions differ | Serious: 178 of 380 events needed | None | HR 0.64 (0.48-0.87) | 63 fewer (94 to 23 fewer) | Very low |
| Cardiovascular death | 2 (RCT) | Not serious | Not serious | Not serious | Very serious | None | RR 0.91 (0.62-1.32) | 4 fewer (14 fewer to 12 more) | Low |
| Stroke | 2 (RCT) | Not serious | Not serious | Not serious | Very serious | None | RR 1.00 (0.56-1.77) | 0 fewer (19 fewer to 33 more) | Low |
| New permanent pacemaker | 1 (RCT) | Not serious | Not applicable | Serious: 1 trial of 1 balloon-expandable valve | Not serious | Approximate denominators | RR ~1.9 (1.3-2.9) | 80 more (28 to 132 more) | Moderate |
| Health status (KCCQ) | 2 (RCT) | Very serious: open label, patient-reported | Not assessable (not pooled) | Not serious | Not serious | None | Not pooled | +9.6 at 1 y; +6.6 at 2 y | Low |
| Death, observational | 7 (cohort) | Very serious: confounding by indication; time-related bias | Not serious ($I^2$ = 8%) | Serious: AVR vs no AVR rather than early vs deferred | Not serious | Inconsistent with randomized evidence | HR 0.52 (0.43-0.64) | — | Very low |

GRADE = Grading of Recommendations Assessment, Development and Evaluation. *The trials enrolled different phenotypes, but the effect did not differ between them. †Both trials measured HF hospitalization, at different time points (1 y in TAVR UNLOAD, 2 y in PROGRESS). Observational evidence started at low certainty. AVR = aortic valve replacement; HF = heart failure; HR = hazard ratio; KCCQ = Kansas City Cardiomyopathy Questionnaire; RCT = randomized controlled trial; RR = risk ratio.

### Supplemental Table 14. Applicability of the Evidence and Ongoing Trials

| Population or question | Randomized evidence | What it shows | Certainty | What it supports |
|---|---|---|---|---|
| Age ≥65 y, predominantly symptomatic (96%), mostly preserved LVEF | PROGRESS (n = 744) | No detectable mortality difference; fewer HF hospitalizations; better health status; more pacemakers | Low (death, HF hospitalization); moderate (pacemaker) | Shared decision on timing |
| HFrEF on guideline-directed therapy | TAVR UNLOAD (n = 178); PROGRESS LVEF <50% subgroup (n = 63) too small to inform | No mortality difference; HF estimates in the same direction but imprecise; better health status at 1 y | Low | Insufficient to recommend early TAVR |
| Advanced cardiac damage (stage 2-4) | PROGRESS subgroup (exploratory) | No evidence that benefit is confined to advanced damage | Very low | Does not support selecting patients by advanced damage |
| Asymptomatic moderate AS | 3.6% of PROGRESS | No informative randomized evidence | — | No evidence for early AVR |
| Surgical AVR; self-expanding valves | None | — | — | No evidence |
| Age <65 y | None (excluded from PROGRESS) | — | — | Lifetime management considerations dominate |
| Ongoing randomized trials | EXPAND TAVR II (NCT05149755; self-expanding valve; n ≈ 650; enrollment closed, primary completion expected September 2027); TIAMAR (NCT05310461; early vs deferred AVR in moderate AS with mitral regurgitation; n ≈ 80; recruiting); NCT07532317 (balloon-expandable Myval TAVR vs guideline-directed therapy, age ≥65 y; n ≈ 778; not yet recruiting) | — | — | — |

AS = aortic stenosis; AVR = aortic valve replacement; HF = heart failure; HFrEF = heart failure with reduced ejection fraction; LVEF = left ventricular ejection fraction; TAVR = transcatheter aortic valve replacement.
